# Key predictors of attending hospital with COVID19: An association study from the COVID Symptom Tracker App in 2,618,948 individuals

**DOI:** 10.1101/2020.04.25.20079251

**Authors:** Mary Ni Lochlainn, Karla A. Lee, Carole H. Sudre, Thomas Varsavsky, M. Jorge Cardoso, Cristina Menni, Ruth C. E. Bowyer, Long H. Nguyen, David A. Drew, Sajaysurya Ganesh, Julien Lavigne du Cadet, Alessia Visconti, Maxim B. Freidin, Marc Modat, Mark S Graham, Joan Capdevila Pujol, Benjamin Murray, Julia S El-Sayed Moustafa, Xinyuan Zhang, Richard Davies, Mario Falchi, Jonathan Wolf, Tim D. Spector, Andrew T. Chan, Sebastien Ourselin, Claire J. Steves, on behalf of the COPE Consortium

## Abstract

**Objectives:** We aimed to identify key demographic risk factors for hospital attendance with COVID-19 infection.

**Design:** Community survey

**Setting:** The COVID Symptom Tracker mobile application co-developed by physicians and scientists at King’s College London, Massachusetts General Hospital, Boston and Zoe Global Limited was launched in the UK and US on 24^th^ and 29^th^ March 2020 respectively. It captured self-reported information related to COVID-19 symptoms and testing.

**Participants:** 2,618,948 users of the COVID Symptom Tracker App. UK (95.7%) and US (4.3%) population. Data cut-off for this analysis was 21^st^ April 2020.

**Main outcome measures:** Visit to hospital and for those who attended hospital, the need for respiratory support in three subgroups (i) self-reported COVID-19 infection with classical symptoms (SR-COVID-19), (ii) selfreported positive COVID-19 test results (T-COVID-19), and (iii) imputed/predicted COVID-19 infection based on symptomatology (I-COVID-19). Multivariate logistic regressions for each outcome and each subgroup were adjusted for age and gender, with sensitivity analyses adjusted for comorbidities. Classical symptoms were defined as high fever and persistent cough for several days.

**Results:** Older age and all comorbidities tested were found to be associated with increased odds of requiring hospital care for COVID-19. Obesity (BMI >30) predicted hospital care in all models, with odds ratios (OR) varying from 1.20 [1.11; 1.31] to 1.40 [1.23; 1.60] across population groups. Pre-existing lung disease and diabetes were consistently found to be associated with hospital visit with a maximum OR of 1.79 [1.64,1.95] and 1.72 [1.27; 2.31]) respectively. Findings were similar when assessing the need for respiratory support, for which age and male gender played an additional role.

**Conclusions:** Being older, obese, diabetic or suffering from pre-existing lung, heart or renal disease placed participants at increased risk of visiting hospital with COVID-19. It is of utmost importance for governments and the scientific and medical communities to work together to find evidence-based means of protecting those deemed most vulnerable from COVID-19.

**Trial registration:** The App Ethics have been approved by KCL ethics Committee REMAS ID 18210, review reference LRS-19/20-18210

## Introduction

As the coronavirus disease 2019 (COVID-19) pandemic escalates and countries struggle to contain the virus, healthcare systems are under increasing pressure as unprecedented numbers require hospitalisation and respiratory support. Emergency departments and intensive care units worldwide are under strain, and medical resources are being diverted to tackle the crisis. There is a pressing need to identify risk factors for severe disease, and particularly to identify key predictors of hospitalisation amongst patients with COVID-19.

In order to address this, we used self-reported data collected on the COVID Symptom Tracker app (1) to identify key demographic risk factors for hospitalisation and the need for respiratory support in the context of COVID-19.

## Materials and Methods

As part of the COronovirus Pandemic Epidemiology (COPE) Consortium (1), the COVID Symptom Tracker smartphone application (“app”) co-developed by King’s College London, Massachusetts General Hospital, and Zoe Global Limited was launched in the UK on 24th March 2020 and was available in the US beginning 29^th^ March 2020 (1,2). Individuals without symptoms are encouraged to use the app. It captured self-reported information related to COVID-19 symptoms. On first use, the app records the user’s self-reported location, age, and core health risk factors (Table 1). At this point height and weight are self-reported, allowing calculation of body mass index (BMI). With continued use, participants provide daily updates on symptoms, information on health care visits, COVID-19 testing results, and whether they are seeking healthcare, including the level of intervention and related outcomes.

**Table 1.**
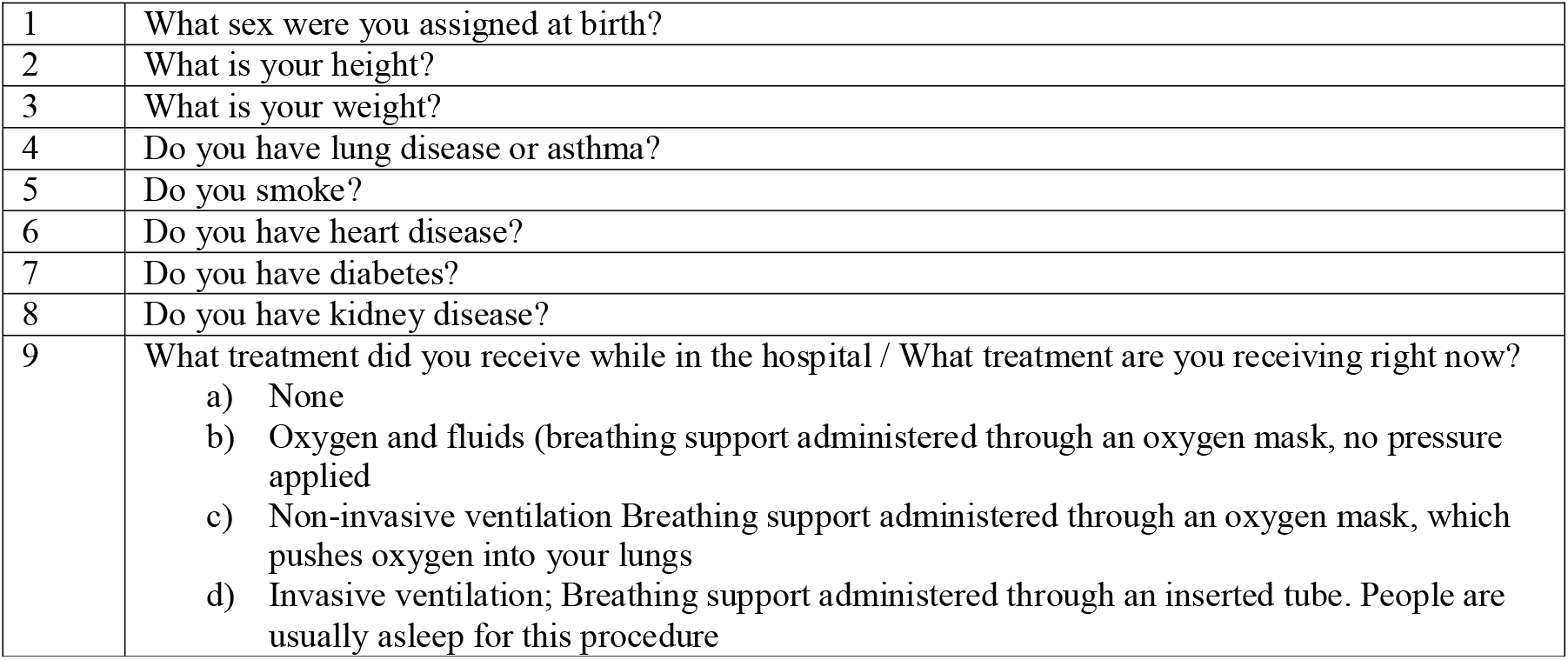
Relevant questions asked in the COVID Symptom Tracker App

|  |  |
| --- | --- |
| 1 | What sex were you assigned at birth? |
| 2 | What is your height? |
| 3 | What is your weight? |
| 4 | Do you have lung disease or asthma? |
| 5 | Do you smoke? |
| 6 | Do you have heart disease? |
| 7 | Do you have diabetes? |
| 8 | Do you have kidney disease? |
| 9 | What treatment did you receive while in the hospital / What treatment are you receiving right now?<br>a) None<br>b) Oxygen and fluids (breathing support administered through an oxygen mask, no pressure applied)<br>c) Non-invasive ventilation Breathing support administered through an oxygen mask, which pushes oxygen into your lungs<br>d) Invasive ventilation; Breathing support administered through an inserted tube. People are usually asleep for this procedure |

### Assessment of exposure, outcomes, and covariates

Exposure, outcome and covariates were all ascertained via the app. A subset of individuals reported being tested for COVID-19. BMI was calculated as kg/m^2^ and considered as a categorical variable (3). Classical symptoms of COVID-19 are defined as “high fever and persistent cough for several days”. This was either directly selfreported as an outcome of an enrolment question or if participants reported jointly fever and persistent cough for more than two days. Visit to hospital was recorded if the location was ever self-recorded as hospital or “back from hospital”.

### Statistical analysis

Data from the app were downloaded and only records where the self-reported characteristics fell within the following ranges were utilised for further analyses: age between 16 (18 in the US) and 100; BMI between 16 and 55 kg/m^2^.

Separate logistic regression models were fit to predict two different outcomes:

1. Visit to hospital as outcome were fit to test for association between i) self-reported obesity and ii) chronic lung disease and asthma, heart disease, diabetes and kidney disease in the following groups: 1) self-reported COVID-19 infection with classical symptoms (SR-COVID19); 2) self-reported positive COVID-19 test results (T-COVID19); 3) imputed/predicted COVID-19 infection based on symptomatology (I-COVID19) Imputation for testing positive for COVID was performed using the data at day of maximum sum of symptoms and applying a logistic regression using coefficients defined previously (2). 100 samples from the coefficient distribution were sampled to create the predictions. A participant was considered as positive 50% of the time. Please see Supplementary Table 1 of the coefficients for the logistic regression.
2. The need for respiratory support (oxygen or ventilation) as the outcome to test for similar association with comorbidities in the specified groups was carried out, for participants who had provided information about the treatment received in hospital.

Sign of recovery was defined as a decrease of more than two points in the sum of reported symptoms compared to the maximum sum of symptoms.

## Results

The total studied sample comprised 2,618,948 individuals who supplied their self-assessment on the COVID Symptom Tracker app after quality control, including 27.4% males. The majority of app users were based in the UK (95.7%), and the remainder in the US (4.3%). Mean age (SD) was 40.3 (13.9) years and mean BMI was 26.74 (5.74). A total of 171,899 (10.5%) replied positively to the question: “Have you already had COVID-19?” 8,4260 (3.2%) replied positively to the question enquiring about classical symptoms of COVID-19.

Diabetes was reported by 75,163 (2.9%), heart disease by 55,196 (2.1%), lung disease by 316,845 (12.1%) and kidney disease by 16,177 (0.6%). 16.3% of the sample reported at least one of the following comorbidities: diabetes, lung, heart or kidney disease. Table 2 summarises the demographic characteristics in the different population groups following the removal of those for whom the outcome (recovery or hospital visit) was still unknown.

**Table 2.**
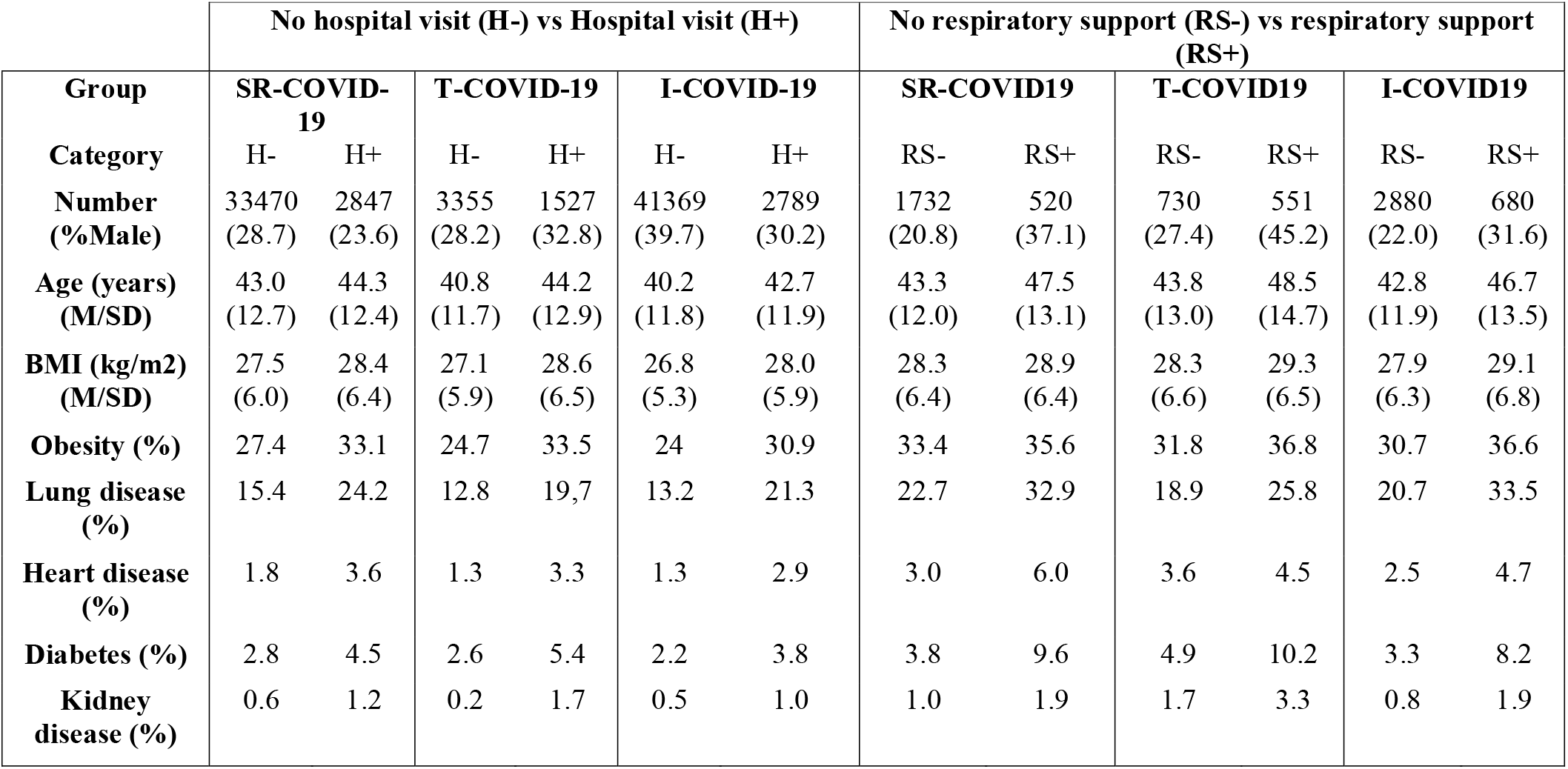
Demographic characteristics of the study population. M = Mean; SD = standard deviation; SR-, self-reported; T-, tested; I-, imputed COVID-19

| Group | No hospital visit (H-) vs Hospital visit (H+) |  |  |  |  |  | No respiratory support (RS-) vs respiratory support (RS+) |  |  |  |  |  |
| --- | --- | --- | --- | --- | --- | --- | --- | --- | --- | --- | --- | --- |
|  | SR-COVID-19 |  | T-COVID-19 |  | I-COVID-19 |  | SR-COVID19 |  | T-COVID19 |  | I-COVID19 |  |
| Category | H- | H+ | H- | H+ | H- | H+ | RS- | RS+ | RS- | RS+ | RS- | RS+ |
| Number | 33470 | 2847 | 3355 | 1527 | 41369 | 2789 | 1732 | 520 | 730 | 551 | 2880 | 680 |
| (%Male) | (28.7) | (23.6) | (28.2) | (32.8) | (39.7) | (30.2) | (20.8) | (37.1) | (27.4) | (45.2) | (22.0) | (31.6) |
| Age (years) | 43.0 | 44.3 | 40.8 | 44.2 | 40.2 | 42.7 | 43.3 | 47.5 | 43.8 | 48.5 | 42.8 | 46.7 |
| (M/SD) | (12.7) | (12.4) | (11.7) | (12.9) | (11.8) | (11.9) | (12.0) | (13.1) | (13.0) | (14.7) | (11.9) | (13.5) |
| BMI (kg/m2) | 27.5 | 28.4 | 27.1 | 28.6 | 26.8 | 28.0 | 28.3 | 28.9 | 28.3 | 29.3 | 27.9 | 29.1 |
| (M/SD) | (6.0) | (6.4) | (5.9) | (6.5) | (5.3) | (5.9) | (6.4) | (6.4) | (6.6) | (6.5) | (6.3) | (6.8) |
| Obesity (%) | 27.4 | 33.1 | 24.7 | 33.5 | 24 | 30.9 | 33.4 | 35.6 | 31.8 | 36.8 | 30.7 | 36.6 |
| Lung disease (%) | 15.4 | 24.2 | 12.8 | 19.7 | 13.2 | 21.3 | 22.7 | 32.9 | 18.9 | 25.8 | 20.7 | 33.5 |
| Heart disease (%) | 1.8 | 3.6 | 1.3 | 3.3 | 1.3 | 2.9 | 3.0 | 6.0 | 3.6 | 4.5 | 2.5 | 4.7 |
| Diabetes (%) | 2.8 | 4.5 | 2.6 | 5.4 | 2.2 | 3.8 | 3.8 | 9.6 | 4.9 | 10.2 | 3.3 | 8.2 |
| Kidney disease (%) | 0.6 | 1.2 | 0.2 | 1.7 | 0.5 | 1.0 | 1.0 | 1.9 | 1.7 | 3.3 | 0.8 | 1.9 |

Across all tested groups, obesity (BMI>30) was associated with a higher risk of attending hospital with COVID-19 symptomatology; odds ratios (OR) ranged from 1.20 [1.11; 1.31] in the SR-COVID-19 group to 1.40 [1.23; 1.60] in the T-COVID-19 group. Lung disease was the most commonly self-reported comorbidity and was found to be a key predictor for hospital visits in all groups with OR varying from 1.68 [1.43, 1.97] for T-COVID-19 to 1.79 [1.64,1.95] for SR-COVID-19.

Heart disease was a significant risk factor in all but the tested group, notably with a stronger effect in the group I-COVID-19 group (1.80 [1.51; 2.16]). The strongest effect of diabetes was observed for the T-COVID-19 group (1.72 [1.27; 2.31]) but a significant effect was seen for all groups. As expected, kidney disease was less frequently reported within this cohort but was nonetheless found to be a significant predictor for a hospital visit in all groups with OR of 1.46[1.02, 2.09], 4.07 [1.97,8.45] and 1.60 [1.29, 2.16] for the SR-COVID-19, T-COVID-19 and I-COVID-19 groups respectively. Figure 1 plots ORs for the considered risk factors, according to the three defined groups.

**Figure 1.**
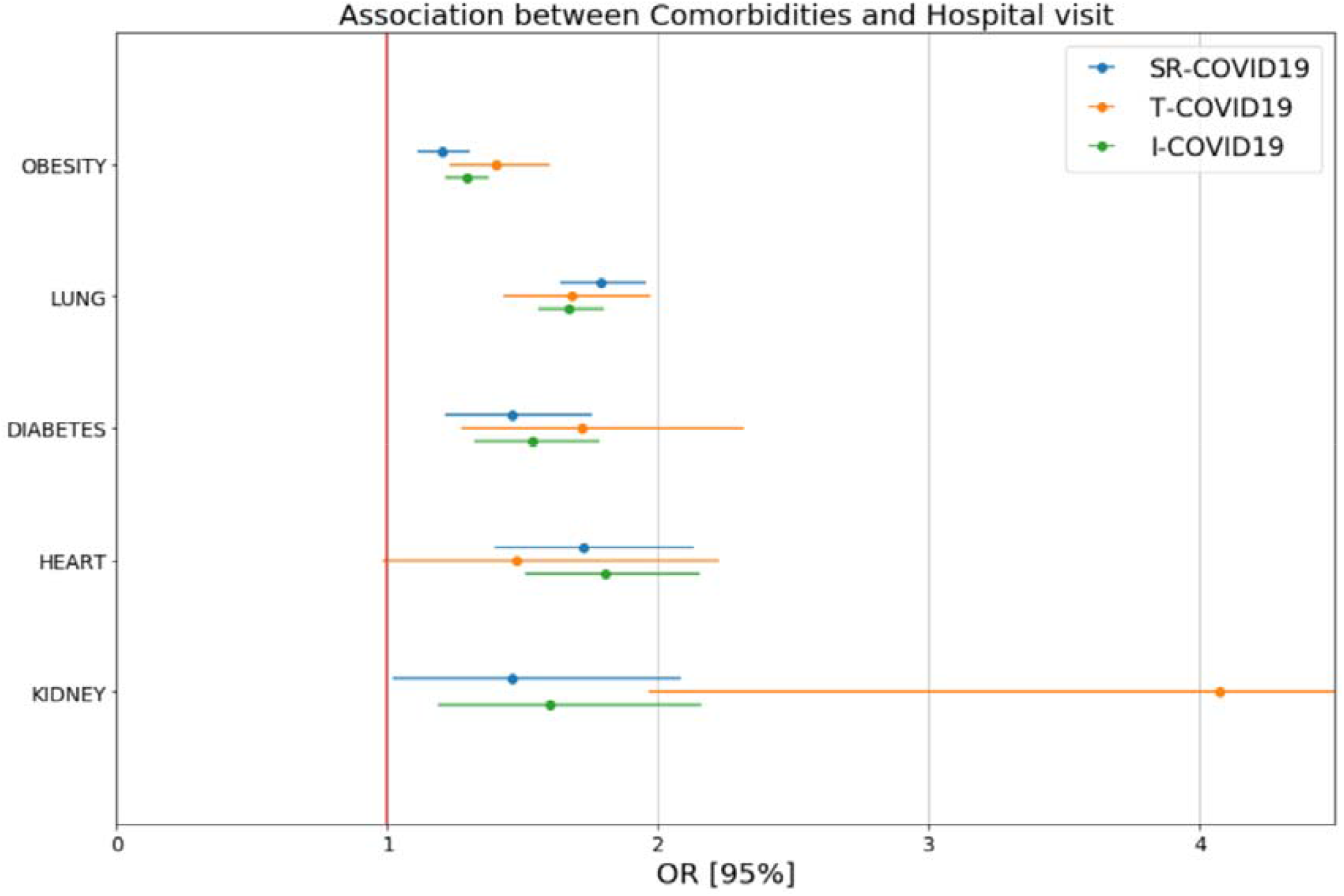
Plotted odds-ratios with their confidence interval for the considered risk factors, according to the three defined groups

While significant in all groups, the effect of age was less strong in SR-COVID-19 (1.007 [1.004 1.01]) when compared with T-COVID-19 (1.02 [1.02;1.03]). Correcting for current smoking did not modify the results but there was a significant difference in smokers’ ratio according to the presented groups with ORs as follows: SR-COVID 1.07 [0.97, 1.17], T-COVID - 1.30 [1.11, 1.52], and I-COVID - 1.16 [1.08 1.25]. Figure 2 illustrates the differrence in age and BMI distribution across the different groups.

**Figure 2:**
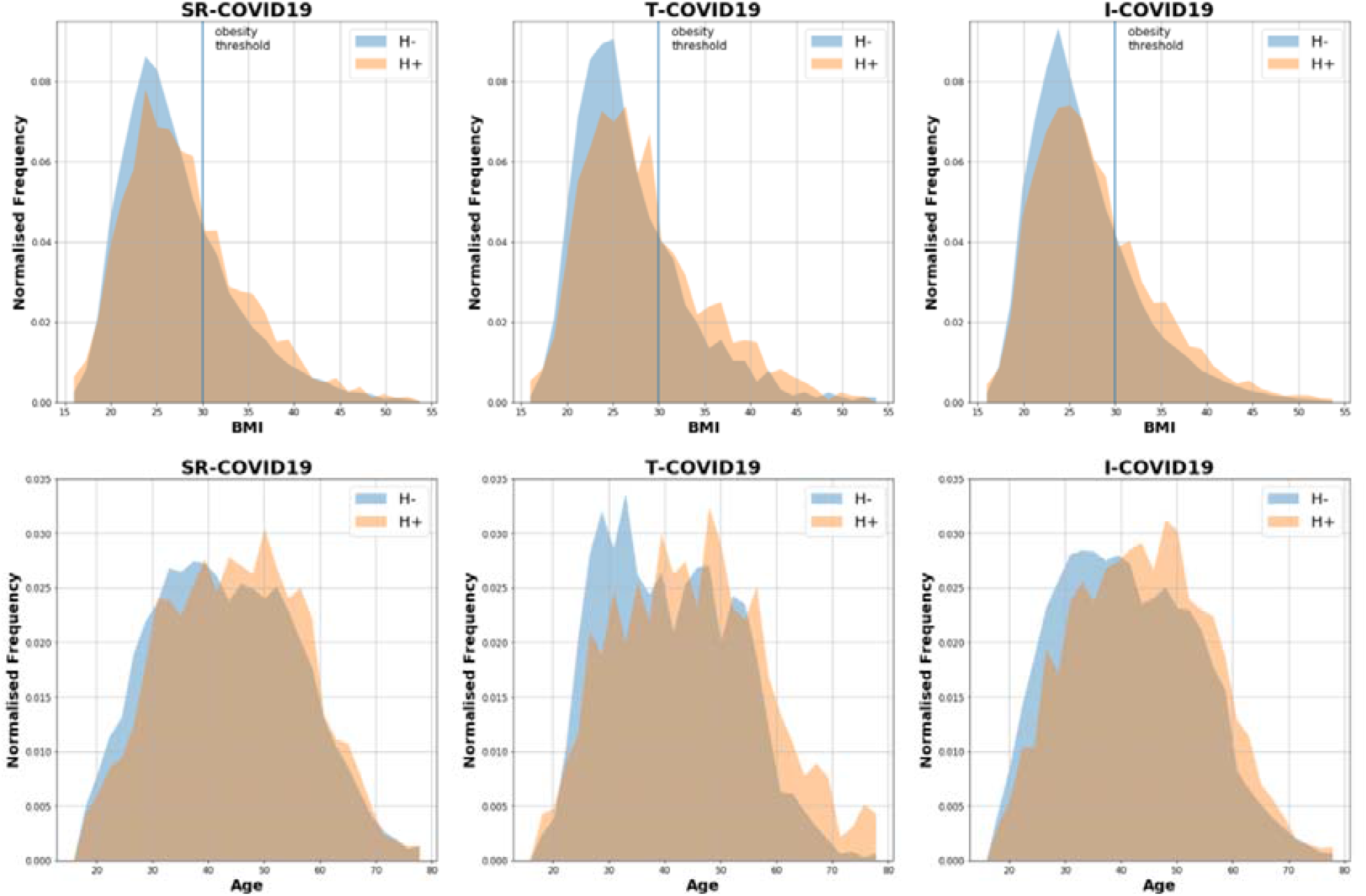
Difference in age and BMI distribution across the different groups H+, hospitalised; H-, not hospitalised; SR-, self-reported; T-, tested; I-, imputed COVID-19

### Requirement for respiratory support (RS)

Amongst those who reported a visit to hospital and then subsequently updated the app with the medical support they received, respiratory support (oxygen and/or ventilation) was reported in 43.0, 23.1 and 19.1% of the cases, for the T-COVID-19, SR-COVID-19 and I-COVID-19 groups respectively. Plotted ORs for needing respiratory support, according to the three defined groups are shown in Figure 3.

**Figure 3:**
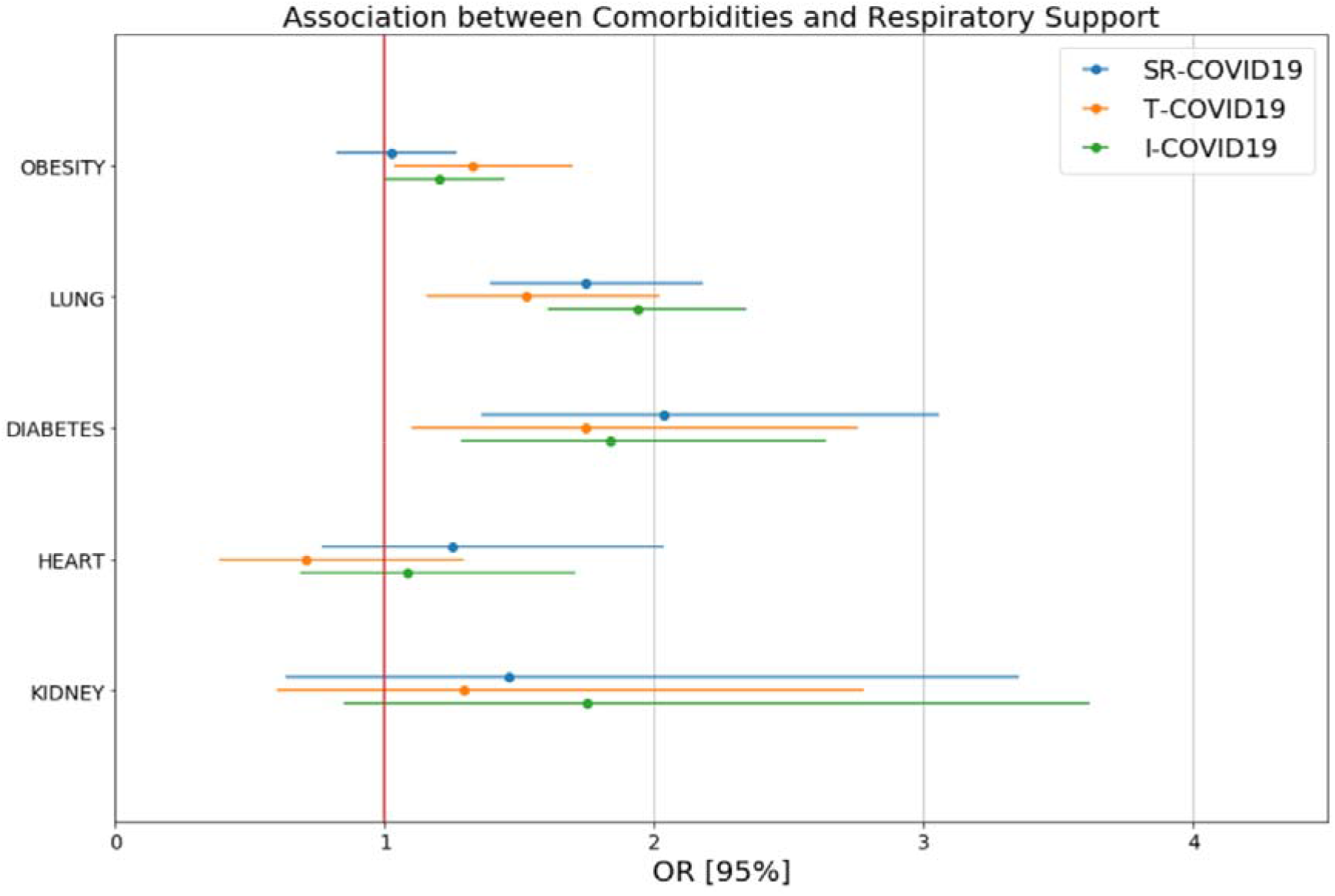
Plotted odds-ratios with their confidence interval for needing respiratory support, according to the three defined groups

Pre-existing lung disease and diabetes were consistently associated with a higher risk of requiring respiratory support with OR ranging from 1.52 for lung disease in T-COVID19 to 2.03 for diabetes in SR-COVID19. Individuals with obesity were notably more likely to require medical support for the T-COVID19 and I-COVID19 groups with OR of 1.33 [1.04 1.70] and 1.20 [1.00,1.44] respectively. Interestingly, while gender was inconsistently associated with hospital visit, male gender was associated with a substantially greater risk for requiring respiratory support, groups with OR of 2.11 [1.65 2.69] and 2.14 [1.72 2.66] and 1.83 [1.28; 2.64] for SR-COVID-19, T-COVID-19 and I-COVID-19 respectively. Age was also more predictive of treatment course than of hospital visit with OR around 1.02 in all three groups.

## Discussion

In this study we found that age, obesity, diabetes and pre-existing lung, renal and cardiac disease, were risk factors for a hospital visit with COVID-19 amongst a large but relatively young, community-based population of app users.

While a number of studies have now reported the increased risk of SARS-CoV-2 in those with diabetes, hypertension and cardiovascular disease (CVD) (4–9), all of which are associated with obesity, the reporting of body mass index (BMI) or obesity has been left out of most initial studies. Only 2 studies have reported BMI in the context of COVID-19 (Table 3). The omission of obesity from initial cohort studies is particularly lamentable when one considers that during the H1N1 outbreak (‘swine flu’), obesity was found to be an independent risk factor for ventilation, as well as increased morbidity and mortality (10,11). Since then, Kwong et al. have published a study that explored the relationship between BMI and seasonal influenza infection using a series of Canada’s cross-sectional population-based health surveys covering twelve influenza seasons (12). Analysis of the retrospective cohort demonstrated that obese people are at greater risk for respiratory hospitalisations during the seasonal flu periods. The World Health Organisation reports that 27.8% of adults in the UK are obese, the third highest rate in Europe. Worldwide, more than 650 million adults are obese (13). Obesity is associated with an increased risk of a wide range of diseases (14), along with being associated with greater risk of infection, and of developing serious complications of common infections (15). Importantly, obese individuals are at higher risk of developing Acute Respiratory Distress Syndrome (ARDS) (16), a common complication of SARS-CoV-2 (5).

**Table 3.** Studies which reported on BMI in the context of COVID-19 infection.

| Study | Participants | Study Methods | BMI Results | Odds ratios/P values |
| --- | --- | --- | --- | --- |
| Peng et al. (49)<br>China | 112 COVID-19 patients with cardiovascular admitted to hospital<br>Critical = 16<br>Non-critical = 96<br>Survivors = 95<br>Non-survivors = 17 | Single-centre, retrospective, observational data<br>Patients divided into critical group and general group according to the severity of the disease and then further divided into non-survivor group and survivor group | BMI of the critical group was significantly higher than that of the general group 25.5 (CI 23.0 - 27.5) kg/m<sup>2</sup> vs. 22.0 (CI 20.0 - 24.0) kg/m<sup>2</sup><br>Among the non-survivors, there were 88.24% of patients had a BMI > 25 kg/m<sup>2</sup>, which was significantly higher than that of survivors (18.95% (18/95)) | Overweight vs healthy weight in critical group: P value =0.003<br>Overweight vs healthy weight in non-survivor group: P value <0.001 |
| Li et al. (20)<br>China | 17 COVID-19 patients<br>5 = discharged<br>12= "quarantined" in hospital | Single-centre retrospective data<br>Comorbidities compared between discharged and quarantined patients | BMI amongst discharged group (24.1 ± 3.5) vs quarantined group (25.3 ± 5.2) | BMI: P value 0.653 |

In the developed world age is now the leading risk factor for many of our most common diseases, including CVD, cancer and neurodegenerative disease (17). Older adults are most affected by annual influenza outbreaks with 75% of influenza-associated deaths in the US in the 2018-2019 season being adults aged >65 years (18). Initial data emerging from the countries first affected by the COVID-19 pandemic reveals that older adults are again disproportionately affected, with multiple studies reporting higher mortality, a more severe disease course and longer length of hospital stay amongst the older population (5,7–9,19,20). The increased vulnerability of older adults to infection is often attributed to ‘immunosenescence’, a term which refers to decline in immunity with advancing age (21). Immunosenescence is associated with increased susceptibility to infection (21), reemergence of latent infections (22), reduced immune surveillance, contributing to diseases such as cancer (23) and the reduced efficacy of vaccines (24,25). Related to this is the concept of ‘inflammageing’, referring to the increase in systemic inflammation noted in older adults (26). Thus, the ageing immune system of an average older adult has higher numbers of circulating inflammatory cells but each with reduced ability to fight infection. However, ageing is a heterogeneous process, and some older adults, especially the physically active have immune systems more comparable to younger adults (27). We show older age as a risk factor for hospitalisation with COVID-19 but also urge more definition and careful consideration by policymakers and healthcare researchers alike. Advice from governments recommending them to stay indoors and “shield” or “cocoon” (28,29), due to their increased risk of morbidity and mortality from COVID-19 may be appropriate on average, but more sophisticated approaches (including immunity testing) should be conducted to avoid blanket decision making on the basis of age alone. The number of people over 60 has doubled since 1980, now exceeding 962 million. Careful consideration must be given to older people in the development of diagnostic, therapeutic, rehabilitation and vaccination programs, in order for such programs to maximise autonomy and minimise the burden on older people both from COVID-19 and the measures introduced to manage it.

Research in other respiratory viruses clearly suggests that patients with chronic respiratory diseases, particularly chronic obstructive pulmonary disease (COPD) and asthma, would be at increased risk of SARS-CoV-2 infection, and of more severe presentations of COVID-19 (30,31). However, chronic lung disease appears to be under-represented in the literature to date for COVID-19 (32); a similar pattern was seen with SARS. With the exception of one single-centre retrospective SARS study (33), clinical studies to date that report on comorbidities in either COVID-19 or SARS, reveal little evidence that chronic lung disease is a key predictor for hospitalisation in these groups (Table 4). To place this table in context, the World Health Organisation (WHO) estimates COPD prevalence to range between 4-20%, depending on the country, and rising considerably with age (34).The lower reported prevalence of chronic lung disease in the literature amongst patients diagnosed with COVID-19 is unexplained. It is possible that, in contrast to the diagnosis of diabetes, there was substantial underdiagnosis or poor recognition of pulmonary disease in patients with COVID-19 patients described in early Chinese cohort studies. Notably, the COVID Symptom Tracker App asked participants: “Do you have lung disease or asthma?”, and we believe that the group answering yes to this question likely includes a large proportion of individuals who self-reported mild asthma (or a previous history of mild asthma). Such diagnoses would possibly not be deemed sufficiently significant for hospital doctors to document as a relevant comorbidity when reporting patients in cohort studies, and so we caution against overinterpretation of the lung disease data herein.

**Table 4.** Studies which reported on pre-existing lung disease in the context of COVID-19 infection.

| Study | Participants | Study Methods | Lung disease reported | Odds ratios/P values (where available) |
| --- | --- | --- | --- | --- |
| Yang et al. (7)<br>China | 52 critically ill adult patients with SARS-CoV-2 pneumonia who were admitted to ICU<br>Non-survivors = 32<br>Survivors = 20 | Single centre retrospective observational study<br>Data were compared between survivors and non-survivors<br>The primary outcome was 28-day mortality | 6% of non-survivors had "chronic pulmonary disease" vs 10% of survivors | Not reported |
| Guan et al. (5)<br>China | 1099 patients with confirmed COVID-19<br>Severe disease = 173 patients<br>Non-severe disease = 926 patients | Data collected from 552 hospitals<br>Retrospective observational study<br>Data compared between severe and non-severe disease (severe = death or requirement for ICU admission/mechanical ventilation) | 0.6% of non-severe cases had COPD vs 3.5% of severe cases | Not reported |
| Zhang et al. (8)<br>China | 140 patients admitted patients with COVID-19<br>Severe disease = 82 patients<br>Non-severe disease = 58 patients | Single centre retrospective study<br>Data compared between severe and non-severe disease (severe = one of the following criteria met: respiratory rate ≥30, O<sub>2</sub> saturations ≤93% at rest and oxygenation index ≤300 mm Hg) | 3.4% of severe disease patients had COPD vs 0% of non-severe cases<br>3.4% of severe disease patients had pulmonary TB vs 0% of non-severe cases | COPD vs not present: p value 0.17<br>TB vs not present: p value 0.17 |
| Zhou et al. (9)<br>China | 191 patients admitted with COVID-19<br>Survivors = 137 patients<br>Non-survivors = 54 died in hospital | Multicentre retrospective study<br>Data compared between survivors and non-survivors<br>Univariable and multivariable logistic regression methods to explore the risk factors associated with in-hospital death. | 7% of non-survivors had COPD vs 1% of survivors | COPD vs not present: Univariable OR 5.4 (0.96-30.4) with p value 0.056 |
| CDC COVID-19 Response Team (50)<br>USA | United States Comorbidity data available on 7,162 (5.8%) for whom COVID-19 reported | US States and territories reporting data directly to Center for Disease Prevention and Control<br>Comorbidity data divided into non-hospitalised, hospitalised non-ICU and ICU<br>"Chronic lung disease" defined as asthma, COPD and emphysema | Of patients needing ICU care, 21% had chronic lung disease<br>Of hospitalised non-ICU patients, 15% had chronic lung disease<br>Of non-hospitalised patients, 7% had chronic lung disease | Not reported |
| Instituto Superiore Di Sanita (44)<br>Italy | Italian Comorbidity data available on 481 patients who died in hospital with COVID-19<br>Non-survivors = 481 | Retrospective, observational data<br>No data on patients who survived | 13.7% of non-survivors had COPD | Not reported |
| Huang et al. (45)<br>China | 41 admitted hospital patients identified as having lab-confirmed COVID-19<br>ICU care = 13<br>Non-ICU care = 28 | Single-centre prospective data collection<br>Outcomes compared between patients who had been admitted to the ICU and those who had not | 8% of ICU care patients had COPD vs 0% of non-ICU patients | COPD vs not present: P value 0.14 |
| Chen et al. (51)<br>China | 99 patients with COVID-19 pneumonia | Single-centre retrospective data | 1% had respiratory system disease | Not reported |
| Liu et al. (52)<br>China | 12 patients with COVID-19 | Single-centre retrospective data | 1 of 12 (8.3%) patients had chronic lung disease | Not reported |
| Wang et al. (53)<br>China | 138 consecutive hospitalised patients with confirmed COVID-19<br>ICU = 36<br>Non-ICU = 102 | Retrospective, single-centre case series<br>Divided into ICU and non-ICU for comparative purposes | 8.3% of ICU patients had COPD vs 1% of non-ICU patients | COPD vs not present: P value 0.054 |
| Wu et al. (54)<br>China | 80 patients with lab-confirmed COVID-19 | Retrospective, multicentre study | 1.25% had a respiratory system disease | Not reported |
| Liu et al. (55)<br>China | 137 COVID-19 infected patients | Retrospective, multicentre study | 1.5% had COPD | Not reported |
| Yang et al. (4)<br>China | 8 studies [including 4 that report respiratory disease and are all included in this table (5,8,45,53)] were included in the meta-analysis, including 46248 patients with COVID-19 | Meta-analysis was to assess the prevalence of comorbidities amongst COVID-19 patients and the risk of underlying diseases in severe patients compared to non-severe patients. | “Respiratory system disease” was found to be the 4<sup>th</sup> most prevalent comorbidity amongst patients | Odds ratio 2±0, (95% CI 1-3%) |
| Xu et al. (56)<br>China | 62 patients admitted with COVID-19 | Retrospective, multicentre study | 2% had COPD | Not reported |
| Emami et al. (6) | Data of 76993 patients presented in 10 articles (including 7 that report respiratory disease and are all included individually in this table(5,45,53–55,57)) with COVID-19 | Meta-analysis to assess the prevalence of comorbidities amongst COVID-19 patients | The incidence rate of COPD in hospitalised COVID-19 patients was 0.95% (95% CI: 0.43%-1.61%) | N/a |
| Booth et al.<br>(58)<br>Canada | 144 adults admitted to 10 hospitals in Toronto, Canada with a diagnosis of suspected/probable SARS in March – April 2003 | Retrospective, observational data<br>Patients not divided into severe, non-severe etc. | 1% of patients had COPD | Not reported |
| Chen et al.<br>(57)<br>Taiwan | 67 patients admitted with SARS in March – July 2003<br>Non-ARDS = 34<br>ARDS = 33 | Single-centre, retrospective, observational data<br>Patients grouped according to whether or not ARDS developed during the clinical course of SARS | 2.9% of non-ARDS patients had COPD or asthma vs 9.1 of ARDS patients | COPD/asthma vs not present:<br>P value 0.356 |
| Ko et al.(33)<br>China | 52 patients admitted with SARS in 2003<br>Survivors = 32<br>Non-survivors = 20 | Single-centre, retrospective, observational data<br>Patients grouped according to survival | 45% non-survivors had “comorbid lung illnesses” vs 0.7% of survivors | “Comorbid lung illnesses” vs not present:<br>P value 0.001 |
| Lee et al.<br>(59)<br>Hong Kong | 138 cases of suspected SARS in March 2003 | Single-centre retrospective, observational data | 2% had “chronic pulmonary disease” | Not reported |

As previously mentioned, many of the initial studies on COVID-19 reported an increased risk of SARS-CoV-2 in those with diabetes, hypertension and CVD (4–9), In the case of diabetes, COVID-19 has also been associated with developing diabetic ketoacidosis, even amongst those who usually have good glycaemic control (35,36). Less well-reported in the context of the pandemic, however, is renal disease. A number of the initial studies from China found that pre-existing renal disease was a risk factor for more severe COVID-19 infection, as summarised in a meta-analysis by Henry and Lippi (37); an overall OR of 3.03 (95% CI 1.09 – 8.47) was found for the association of chronic kidney disease to severe COVID-19 across 4 early studies. In addition, it is becoming apparent that COVID-19 can cause acute kidney injury (AKI) in COVID-19, and this confers a worse prognosis: a Chinese prospective cohort study (n=701) reported that 44% and 27% of COVID-19 patients had proteinuria and haematuria on admission, respectively, with 5.1% of this cohort developed AKI during their hospital stay (38). Importantly, this study also reported higher mortality from COVID-19 for those who had pre-existing kidney disease. As we learn more about AKI in COVID-19, it is becoming paramount to disentangle whether pre-existing renal disease predisposes patients to significant AKI in COVID-19, as one might expect. While we did find a significantly increased risk of attending hospital for those with renal disease, relatively few reported this condition in our cohort (Table 2), which may represent a problem with self-reporting bias. More research is needed to examine the effect of pre-existing renal disease on COVID-19 infection.

Despite the literature consistently showing males to be at higher risk for more severe COVID-19 (7,39,40), we did not find gender assigned at birth to be a predictor of attending a hospital in this study. However, male gender was associated with a substantially greater risk of requiring respiratory support, in all groups. Men have been shown to be less likely to utilise medical services than women (41) and large studies have consistently shown women to report greater numbers of physical symptoms than men (41,42). Such health behaviours may lead to men having a higher threshold for visiting the hospital with COVID-19 than women, along perhaps with less awareness of early symptoms. This may result in males being more unwell when they do seek medical help and contribute somewhat to the stark difference in mortality between genders (43–45).

Our sample size is the largest reported to date, adding significant strength to many predictors of hospitalisation or severe COVID-19 that have been identified in smaller cohort studies to date. We hope that our data will guide clinicians to protect their at-risk patient cohorts and policymakers to consider these vulnerable groups when planning and allocating resource, both during this pandemic and for future similar eventualities. Clearly many questions remain, and the mechanisms by which a number of these predictors of hospitalisation are unclear; much research is needed to define these questions, and quickly.

Our study has a number of limitations. First, all the data collected is self-reported, and questions on comorbidities were somewhat simplified to ease reporting at large scale on an app. Both symptoms and test results may be subject to reporting bias. Secondly, the sampling using an app will under-represent individuals without smartphone devices, including older participants, and is likely to under-represent those severely affected by the disease. Additionally, we are reporting visits, rather than admissions, to hospital; we do not know how many visits resulted in an inpatient stay. While we believe that our sampling provides useful information about the risk of most symptomatic infection, it will not provide insight into very severe disease as the most unwell patients may not record hospitalisation due to incapacitation or even death. Additionally, COVID-19 diagnoses, where confirmed by testing, were likely to be based on RT-PCR which is thought to be between 66-80% sensitive for a single test (46,47). Another important caveat of note is that the individuals on which the model was trained are highly selected because COVID-19 tests are not performed at random (48). The participants were tested because they either displayed severe symptoms, were in contact with COVID-19 positive individuals, were healthcare workers or had travelled to an area of particular risk. Additionally, the app captured whether participants had been diagnosed with COVID-19 but did not specifically ask when. Given that symptoms are recorded at the time of data entry, it is possible that some individuals may no longer have been symptomatic from the virus.

## Conclusion

The key predictors of hospitalisation in the context of COVID-19 affect many in our society. Lung disease, diabetes, heart disease, advanced age and increased BMI were associated with risk of hospitalisation with COVID. Careful planning of the use of immune testing and contact tracing could be particularly relevant for people in these higher-risk groups. This may help minimise the risk that patients living with comorbidities and older people are disproportionately isolated in the months to come. Finally, the presence of these factors should be regarded as an important factor in future risk stratification models for COVID-19. It is of utmost importance for governments and the scientific and medical communities to work together to find evidence-based means of protecting those most vulnerable from COVID-19 in a way which minimises their risk of economic, mental and social implications of isolation.

## Ethics

In the UK, app Ethics has been approved by KCL ethics Committee REMAS ID 18210, review reference LRS-19/20-18210. In the US, the informed consent process was approved by the Partners Human Research Committee (Protocol 2020P000909). All subscribers provided informed consent before submitting responses. The study was registered with clinicaltrials.gov as NCT04331509.

## Data sharing

Data used in this study is available to bona fide researchers through UK Health Data Research using the following link https://healthdatagateway.org/detail/9b604483-9cdc-41b2-b82c-14ee3dd705f6

## Contribution Statement

MNL, KAL, and CHS carried out the analyses and wrote the original draft. MNL, KAL, CHS, TV, MCJ, CM, RCEB, LHN, DAD, AV, MF, MM, MSG, BM, JESM, XZ, MF, TDS, SO, CJS and ATC contributed to conceptualisation, data curation, investigation, methodology, validation and reviewing and editing the draft. SG, JLdC, JC, RD, and JW contributed to the data curation, methodology, resources and software. SO and CJS supervised the project.

## Dissemination Declaration

dissemination of results to all app users is not possible, however results will be shared on our website https://covid.joinzoe.com/, and via our social media, as well as through national and international media channels.

